# Pupil symmetry and Swinging Light Test Performed with VR Headset for Detection of Anisocoria and Relative Afferent Pupillary Defect

**DOI:** 10.64898/2026.09.02.26361629

**Authors:** Maja Štrumbelj, Nataša Vidovič Valentinčič, Ana Fakin

## Abstract

**Purpose:** Pupil examination is a routine examination of patients presenting with ophthalmological or neurological symptoms. Our purpose was to determine the sensitivity of pupillometry using HP Reverb G2 Omnicept virtual reality (VR) headset for detecting anisocoria and relative afferent pupillary defect (RAPD).

**Methods:** Twenty-nine patients were included; Group I with asymmetric optic neuropathy (N=21) and Group II with suspected pathological anisocoria (N=8); alongside 19 controls. VR pupillometry consisted of Pupil symmetry test and Swinging light test. Pathological thresholds were established using upper limits of controls. Results were compared with routine pupil exam. Correlations with functional ophthalmological test results were analyzed.

**Results:** RAPD was detected in 16/21 Group 1 patients using both VR pupillometry and standard exam. Remaining 5 had relative afferent pupillary asymmetry (RAPA) within the control range (0.1–0.2 mm). Significant correlation was found between RAPA/RAPD and inter-eye difference in visual acuity and visual field defect (MD) (R=0.58 and 0.68, p<0.01). All patients with >5,8 dB inter-eye MD difference had RAPD. In Group II, pathological anisocoria (>0.8 mm) was detected using VR pupillometry and 1/8 on standard exam. Qualitative differences in pupillary constriction were observed. In control group anisocoria (0.4–0.8 mm) was detected in 37% (7/19); in all greater in the dark (mean 0.2 mm).

**Conclusion:** In comparison to conventional exam VR pupillometry matched detection rate of RAPD and surpassed detection rate of pathological anisocoria. Longitudinal data on pupillary constriction characteristics could significantly contribute to patient diagnosis and monitoring in clinical studies as well as understanding physiological variability.

## Introduction

Pupil examination is an important part of the clinical assessment of patients with ophthalmological or neurological symptoms and is routinely performed by ophthalmologists, neurologists and emergency care physicians. Asymmetry in pupil size (anisocoria) may indicate damage to the oculomotor nerve or sympathetic nervous system, while an impaired pupillary light reflex (afferent pupillary defect) is an important sign of visual pathway damage, especially in poorly cooperative patients. Pupil measurements objectively define the function of the afferent visual pathway and the balance between the sympathetic and parasympathetic nervous systems; therefore, methods that enable precise assessment of pupil size and pupillary reactions are additionally interesting for research purposes. The aim of this study was to define a new method of pupillometry using virtual reality (VR) headset HP Reverb G2 Omnicept Edition with built-in biometric sensors.

Anisocoria is a condition in which the pupils of the right and left eye are of different sizes (Kılınç Hekimsoy et al. 2022). It is defined as a difference in pupil diameter between the left and right eye of at least 0.4 mm, as such a difference is considered easily detectable with the naked eye (Lam, Thompson & Corbett 1987, Loewenfeld 1977). Anisocoria may be physiological, iatrogenic, or due to pathological conditions, such as oculomotor nerve palsy, Horner syndrome, Adie’s pupil, and local iris injury (Fakin 2023). Physiological anisocoria is a benign pupillary inequality that occurs in healthy individuals without neurological or ocular pathology (Kılınç Hekimsoy et al. 2022, Lam, Thompson & Corbett 1987). Physiological anisocoria is characterized by a small difference between the pupils, usually up to 1 mm, which remains the same in light and darkness. The pupils react well to light and accommodation and rapidly redilate after constriction (‘Anisocoria - EyeWiki’ n.d., Fakin 2023). It is the most common cause of anisocoria and occurs in approximately 20% of people (Lam, Thompson & Corbett 1987, Loewenfeld 1977). Some studies have found that the prevalence of physiological anisocoria depends on light intensity, defined as cd/m2. The prevalence is lower under photopic lighting conditions (10–100 cd/m2) and higher under mesopic lighting conditions (0.1–1 cd/m2) (Kılınç Hekimsoy et al. 2022). Benign alternating anisocoria is a condition in which the larger pupil alternates between the right and left eye within physiological limits (Bremner, Booth & Smith 2004).

An afferent pupillary defect (APD) is a reduced pupillary response to illumination due to decreased light perception at the level of the retina or optic nerve, or due to damage to the visual pathway, anterior to the lateral geniculate nucleus. It is divided into absolute APD, which occurs with complete interruption of afferent sensation and is characterized by a completely absent pupillary constriction to direct illumination of the affected eye, and relative APD (RAPD), in which there is a partial (relative) reduction in the pupillary response on the affected side to direct illumination compared with the response to indirect illumination.

The standard clinical method for detecting RAPD is the swinging flashlight test, which is an important method in clinical practice for detecting partial or asymmetric damage to the afferent pathway. The test is performed in a dimly lit room using a directed light source. The patient looks at a distant fixation target to prevent activation of the accommodation reflex. The light is directed into one eye for approximately 3 seconds, then rapidly moved to the other eye, and the procedure is repeated several times. Illumination of either eye causes symmetrical miosis of both pupils. In RAPD, however, relative dilation occurs when the light is moved to the affected eye (so-called paradoxical dilation). Despite its simplicity, the test is very important in clinical assessment, as it enables rapid recognition of asymmetric retinal or optic nerve disease (Broadway 2012). Depending on the degree of afferent impairment, different grades of RAPD can be detected. Clinically, RAPD assessment is most used in patients with optic neuropathies, such as optic neuritis, anterior and posterior ischemic optic neuropathy, compressive optic neuropathy, and compartment syndrome. RAPD is especially important in posterior optic neuropathies, in which the fundus may appear completely normal, and RAPD may be the only sign in clinical examination that objectively demonstrates a pathological condition (Broadway 2012). The degree of RAPD is usually determined subjectively according to the characteristics of pupillary dilation when the light is moved from the healthy to the affected eye (Broadway 2012). For precise quantification, especially for research purposes, neutral density (ND) filters of different densities have been used in the past (Thompson, Corbett & Cox 1981).

The first use of dynamic infrared pupillometry, which enables accurate pupil measurement in dark, was published in 1958 (LOWENSTEIN & LOEWENFELD 1958), but pupillometers were traditionally custom-made for research purposes and had only limited automation for data acquisition and analysis. In recent years, research approaches have emerged using infrared sensors integrated into different headsets, including virtual reality (VR) headsets. The use of VR technology has several potential advantages: standardized illumination, automated stimulation of each eye separately, simultaneous binocular measurement, and reduced inter-observer variability (Negi et al. 2024). Most studies using VR pupillometry focused either on detection of RAPD or anisocoria, while a comprehensive test detecting both had not been put in routine clinical or research practice.

The aim of the study was to define the sensitivity of automated pupillometry with HP Reverb G2 Omnicept Edition virtual reality headsets, referred to in this study in short as “VR pupillometry”, for determining anisocoria and relative afferent pupillary defect. In addition, we aimed to characterize the features of physiological anisocoria more precisely.

## Methods

This prospective clinical study was conducted in the Virtual Reality Laboratory within the National Center for Comprehensive Rehabilitation of the Blind and Visually Impaired at the Eye Hospital of the University Medical Centre (UMC) Ljubljana. All examinations were performed in accordance with the Declaration of Helsinki on biomedical research involving humans. The study was approved by the Republic of Slovenia National Medical Ethics Committee (decision No. 0120-644/2025-2711-3, application dated 19 December 2025, approved 23 January 2026; Appendix 1). Participants received an oral invitation with an explanation of the purpose, benefits, safety, and course of the study, and confirmed participation by signing written informed consent (Appendix 2). The study used computer equipment, virtual reality headsets and software purchased as part of the ARRS J3-1750 research project and the UMC Ljubljana tertiary project (ID 20220038) “Development of a virtual reality laboratory”. Authors have no conflict of interest.

### Inclusion and exclusion criteria

Healthy participants and patients treated at the Eye Hospital, UMC Ljubljana who met the inclusion criteria below were enrolled in the prospective study. Patients were identified among those followed in the Neuro-ophthalmology Clinic based on ophthalmological examination findings and routine tests, including pupil examination. Patients with neurological impairments were excluded because of their possible influence on pupillary reactions and study cooperation. For the control group, we invited healthy individuals, including patient companions, Eye Clinic employees, and relatives, who had no ocular or neurological disease. All participants were ≥ 18 years old.

Three groups of participants were included in the study:

I. Patients with unilateral or bilateral optic neuropathy (anterior ischemic neuropathy, optic neuritis, pituitary tumor). This group included 21 participants, 6 men and 15 women, mean age 58 years, range 23–90 years.
II. Patients with anisocoria. This group included 8 participants, 5 men and 3 women, mean age 58 years, range 25–73 years.
III. Control group. This group included 19 participants, 5 men and 14 women, mean age 45 years, range 23–83 years.

### VR pupillometry

Participants underwent measurements of pupil size in dark and light using VR headset HP Reverb G2 Omnicept Edition (Hewlett Packard, Spring, TX, USA), which contains an integrated sensor for measuring pupil size, and ability to display different images or illumination to each eye. IC pupillometry version 2.5 software (Synthesius, Ljubljana, Slovenia) was used to conduct the testing and display the acquired data. Eye illumination intensity during the test was the maximum headset screen illumination, 150 cd/m2 (‘HP Reverb G2 vs Pimax Crystal’ 2024).

Participants completed two tests: 1) Pupil symmetry test, which allows assessment of anisocoria by measuring right and left pupil size in light and dark. In the headset, darkness is first shown for 10 s, then both eyes are illuminated for 5 s, followed again by bilateral darkness. This is repeated alternately four times. 2) Swinging light test, which allows assessment of RAPD. Darkness is first shown for 10 s. The right and left eye are then illuminated alternately, each for 3 s. The right eye is illuminated first. During the examination, the headset continuously records pupil diameter. Measurement is absent during blinks. At the end of the examination, the computer software plots the trace demonstrating of pupil diameter for each eye separately (Fig. 1).

**Figure 1:**
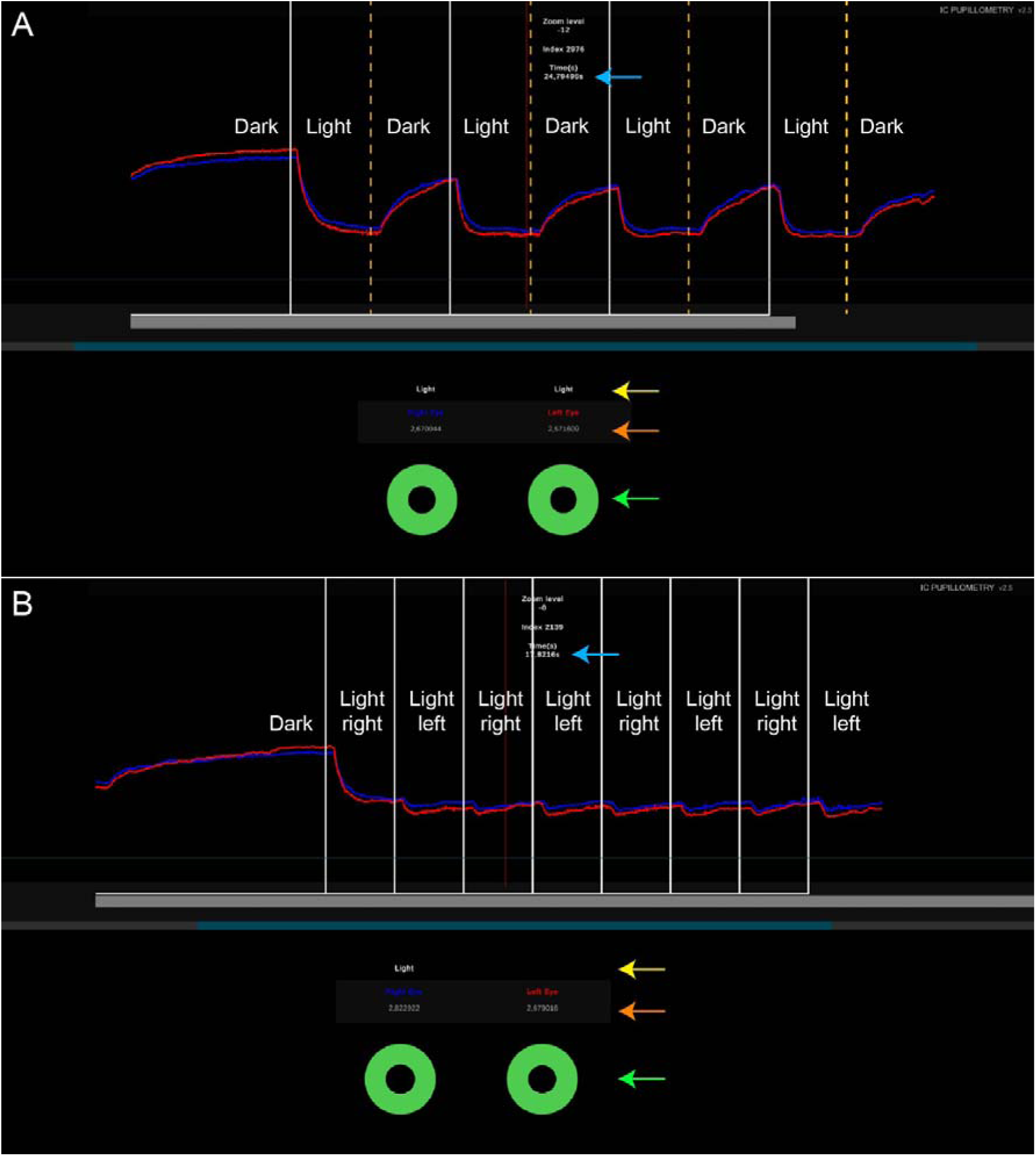
Example of a longitudinal recording of the Pupil symmetry test (A) and Swinging light test (B). The X-axis represents time and the Y-axis pupil diameter. Changes in right-pupil diameter are shown in blue and changes in left-pupil diameter in red. The timer is shown in the upper part (blue arrow). White vertical lines show time points when the light was switched on, vertical orange dashed lines show time points when the light is switched off and red vertical line shows the manually selected time point. For the selected time point, schematic representation of pupil diameters is shown below (green arrow), as well as the exact diameter of each pupil in mm (orange arrow) and illumination conditions (light on/off) (yellow arrow).

From the longitudinal VR pupillometry recordings, data on the diameter of the left and right pupils in mm were obtained and recorded at selected reading points. The reading points were selected immediately before each change in lighting condition (red line on Fig. 1A), allowing maximal adaptation to the lighting condition. If a blink occurred in the interval in which the measurement should have been read, the measurement was read immediately before the blink. Measurement was performed only if there was reliable trace in the 0,1s time window before the change in lighting conditions. For statistical purposes the pupil diameter was rounded to 0.1 mm to account for the limited accuracy of pupillometers, estimated at 0.03–0.1 mm (Hsu & Kuo 2023). For the Pupil symmetry test, repeated measurements of each eye for each light condition (darkness and bilateral illumination) were averaged. Using these averages, we calculated the inter-eye difference (asymmetry) during darkness and on illumination for each participant. For the Swinging light test, repeated measurements of each eye during for each light condition (right- and left-eye illumination) were averaged. Using these averages, we calculated the difference between the pupillary diameters of each eye during right- and left-eye illumination (direct and indirect response). Finally, we calculated the mean of these values, which we termed relative afferent pupillary asymmetry (RAPA), or relative afferent pupillary defect (RAPD), if it exceeded normal range. In patients RAPA/RAPD was also determined for the eye on the affected side (RAPAa/RAPDa), imitating the observation during manual testing. Example of calculations is shown in Table 1.

**Table 1.** Example of analysis of VR Swinging light test results.

| Pupil diameter (mean of 4 repeated measurements) [mm] |  |  |  | Difference between direct and indirect response [mm] |  | Mean difference from previous two columns [mm] (RAPA/RAPD) |
| --- | --- | --- | --- | --- | --- | --- |
| Illuminated RE |  | Illuminated LE |  |  |  |  |
| RE (direct response) | LE (indirect response) | RE (indirect response) | LE (direct response) | RE | LE |  |
| 4.5 | 4.3 | 2.6 | 2.5 | 1.9 (RAPDa) | 1.8 | 1.9 |

This participant had right-sided optic neuropathy. RE – right eye, LE – left eye. Note larger pupil diameter on both eyes (direct and indirect response) on RE illumination.

### Clinical assessment

Patients with optic neuropathy (Group 1) underwent routine swinging light exam by a senior specialist of ophthalmology using penlight to alternately illuminate each eye for 3 seconds. RAPD was classified as reliably positive, reliably negative, and not reliably determinable. All participants were photographed under high and low illumination (lights on and off in a dim room) using portable camera (iPhone SE). During dim-light photography, the light capture was increased using longer exposure. Photographs were imported into Fiji software [available at https://fiji.sc/, 1 July 2026] and pupil diameters measured with measuring tool calibrated to the horizontal corneal diameter of each eye, assuming that (“white-to-white diameter”) is approximately 12 mm in adults (Rüfer, Schröder & Erb 2005). Inter-eye difference in pupil diameter under both lighting conditions was calculated. Data on best corrected Snellen visual acuity (BCVA), and static perimetry performed using Octopus G2 top protocol (Haag-Streit, Switzerland) was collected from the medical records. For statistical analysis, Snellen visual acuity was converted to logMAR (Bailey & Lovie 1976). From the visual-field reports, we recorded the mean defect (MD) in dB for each eye separately, where 0 dB represents normal sensitivity (device norm). If visual-field testing was not possible to perform due to severe visual loss, we assumed that patients would not see any stimulus during the examination, corresponding to a defect of 27.5 dB.

### Statistical analysis

Statistical analysis was performed using IBM SPSS Statistics, version 27 (IBM Corp., Armonk, NY, USA). P value of < 0.05 was used as the threshold for statistical significance. Normality of distribution was assessed with the Shapiro-Wilk test. Normally distributed variables were described with the mean and non-normally distributed variables with the median. Because of the small patient groups, ranges were reported. Associations between continuous variables (RAPA/RAPD grade, BCVA, MD) were assessed with Pearson correlation.

## Results

VR Swinging light test revealed no RAPA in 16% (3/19), RAPA of 0.1 mm in 79% (15/19), and RAPA of 0.2 mm in 5% (1/19) controls; the average being 0,1 mm. The upper limit (>0.2 mm) was determined as the threshold for positive RAPD on VR pupillometry. Using the threshold of >0,2 mm, positive RAPD was identified in 16/21 patients with optic neuropathy and a subthreshold RAPA in the remaining 5/21 patients. The RAPA / RAPD grade determined by measuring only the affected side were comparable and did not change the percentage of positive RAPD (Table 2). Results of VR pupillometry matched clinical RAPD assessment. Positive RAPD on VR pupillometry was found in all patients (16/16) with clinically reliably positive RAPD and in none with reliably negative or not reliably determinable RAPD. The degree of RAPA was 0.4–1.9 mm in the group with reliably positive RAPD, 0.1 mm in both patients with reliably negative RAPD, and 0.1–0.2 mm in patients with not reliably determinable RAPD. VR recordings and visual fields of representative cases are shown in Figures 2–5. Patient 21 with right-sided optic neuritis underwent repeated VR pupillometry after two months, when visual function had improved (Table 2).

**Figure 2.**
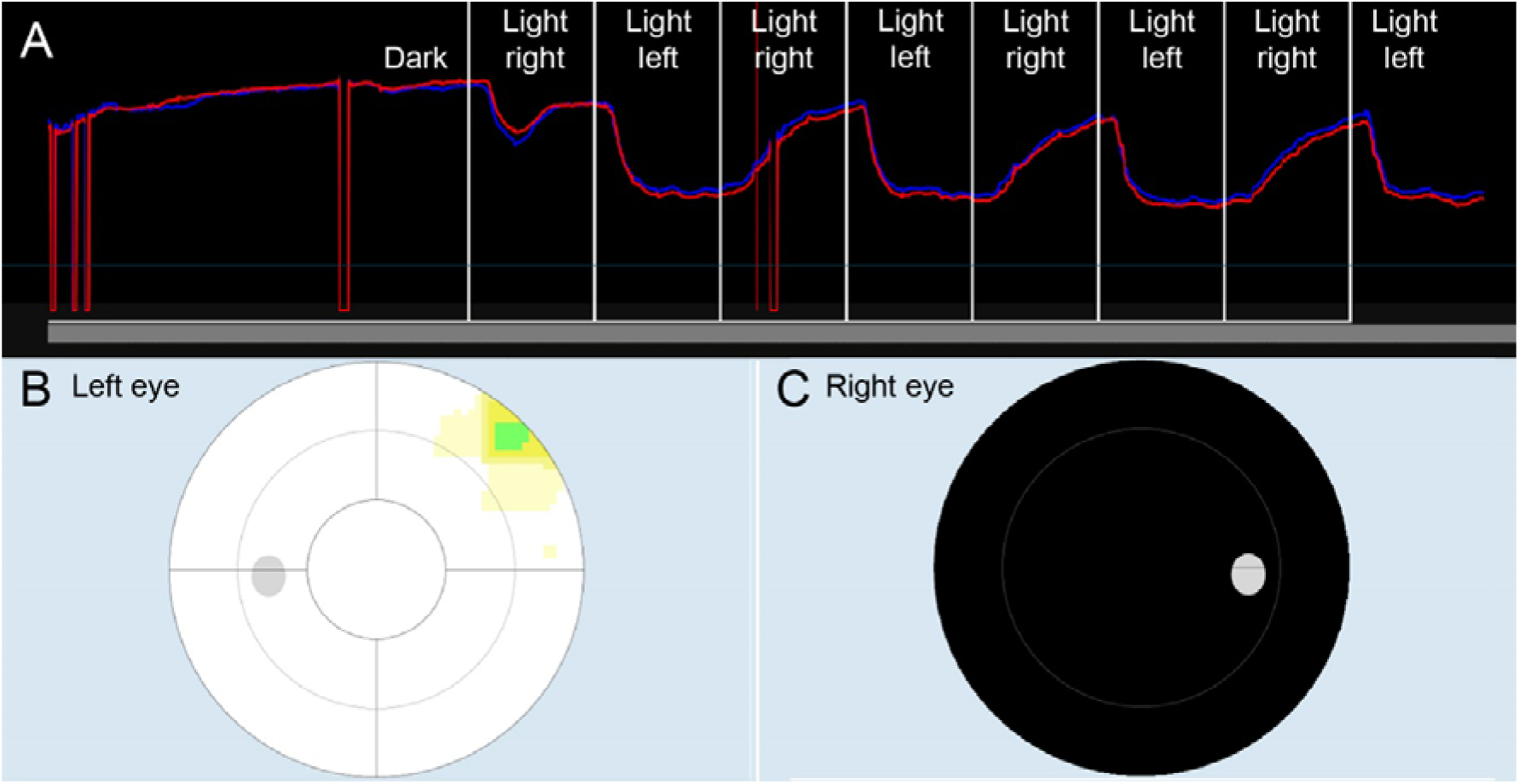
Example of positive RAPD. VR Swinging light test (A) and static perimetry of left (B) and right (C) eye of patient 39 with right-sided optic neuritis. The patient had a positive RAPD on VR pupillometry in the magnitude of 1,9 mm, as well as a clinically positive RAPD. BCVA was hand movements (2.5 logMAR) in the right eye and 1 (0 logMAR) in the left eye.

**Figure 3.**
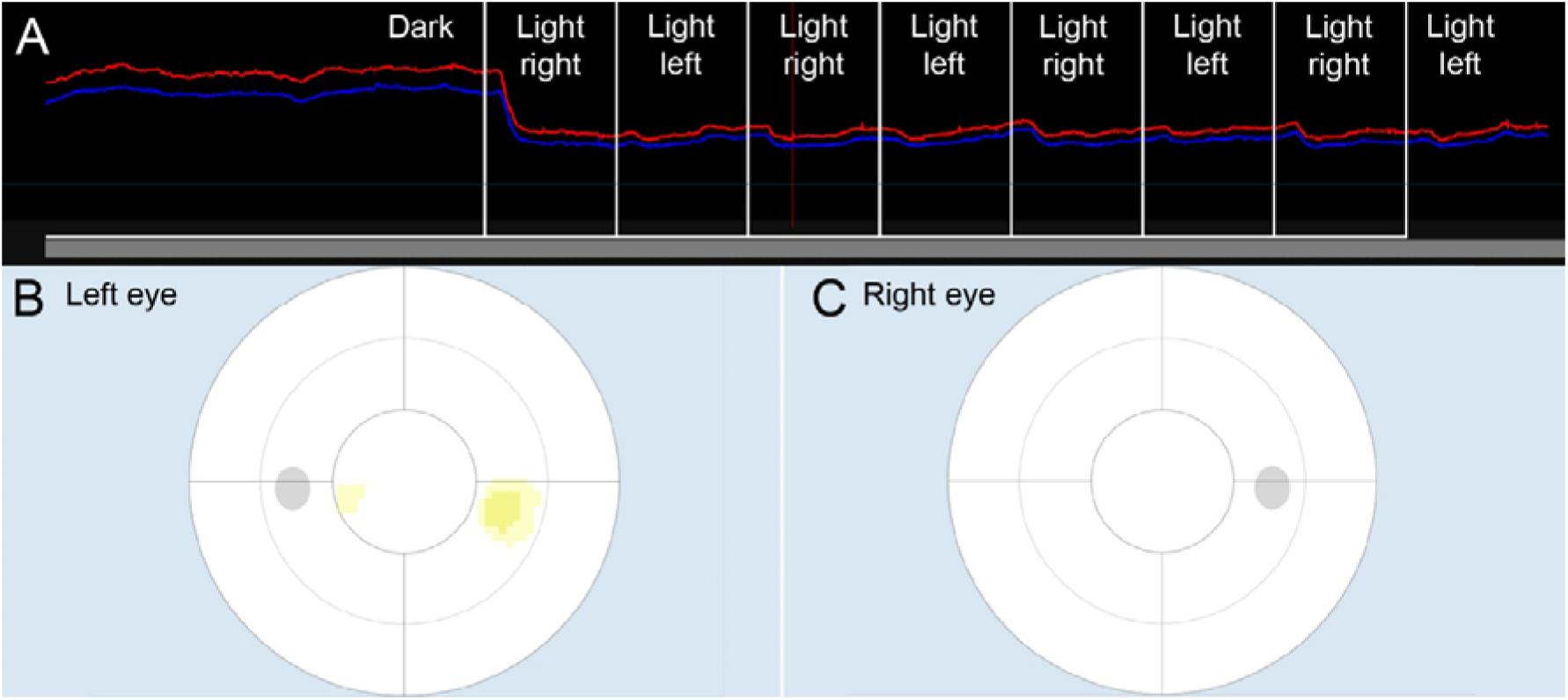
Example of negative RAPD in patient 26 after recovery from left-sided optic neuritis. VR Swinging light test (A) and static perimetry of left (B) and right (C) eye are shown. The patient had a subthreshold RAPA on VR pupillometry (A) in the magnitude of 0,1 mm and clinically negative RAPD. BCVA was 1,0 (0 logMAR) bilaterally.

**Figure 4.**
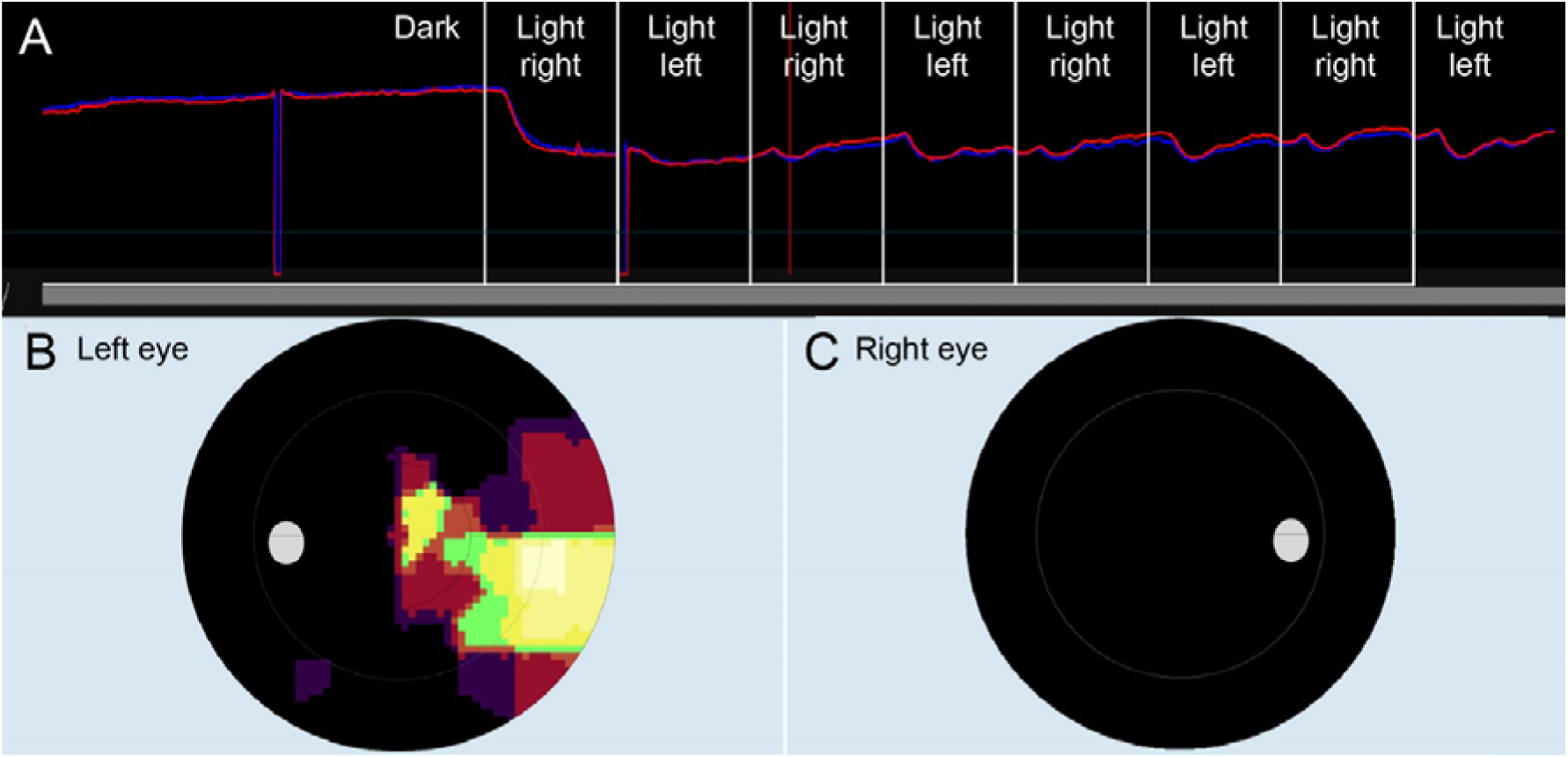
Example of negative RAPD in patient 27 with severe bilateral visual loss due to N- AION. VR Swinging light test (A) and static perimetry of left (B) and right (C) eye are shown. The patient had a subthreshold RAPA on VR pupillometry in the magnitude of 0,1 mm and clinically not reliably determinable RAPD. BCVA was light perception (2.8 logMAR) on the right and 0.06 (1.2 logMAR) on the left eye.

**Figure 5.**
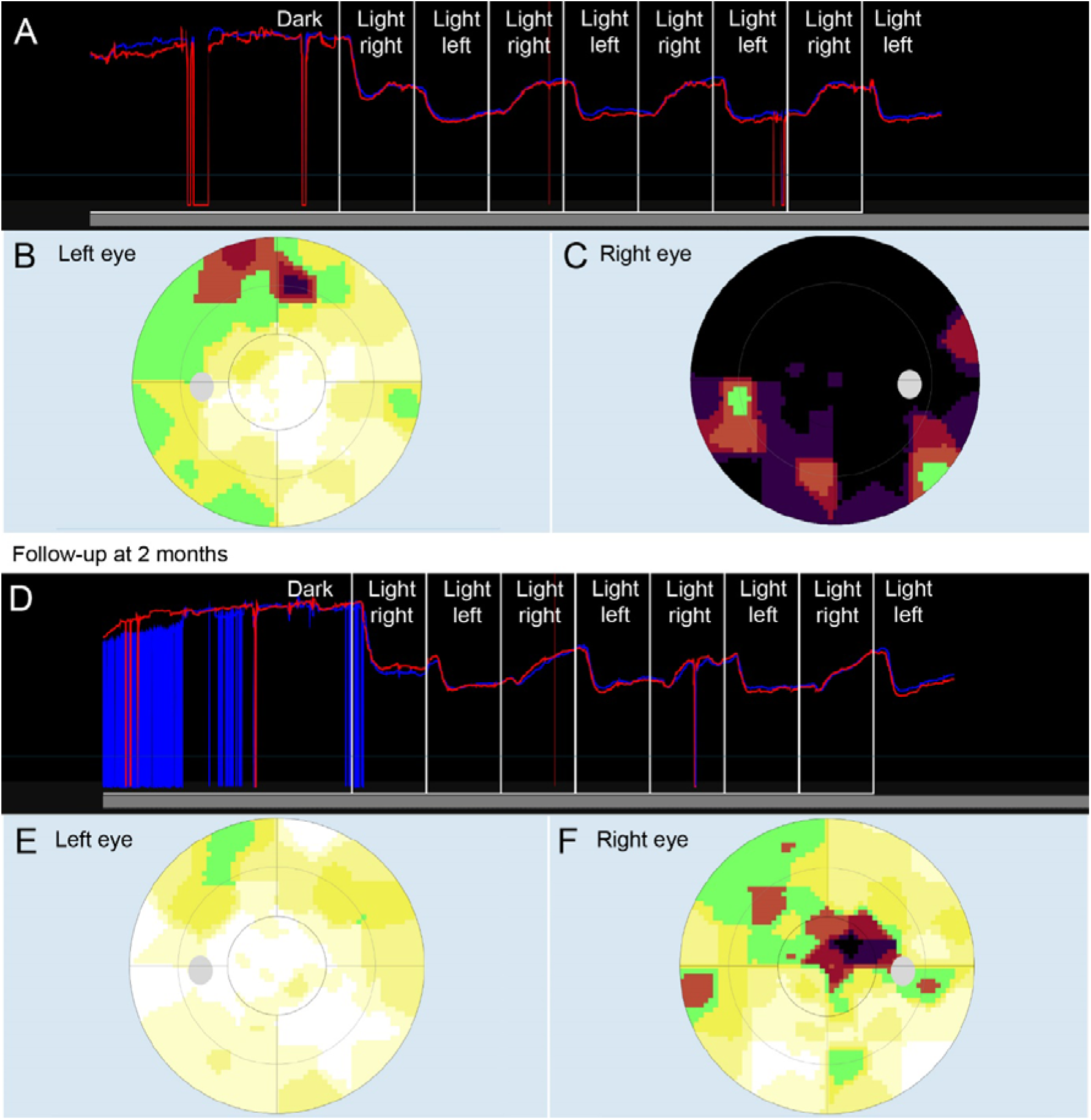
VR Swinging light test (A) and static perimetry of left (B) and right (C) eye of patient 21 with right-sided optic neuritis at presentation and at two-month follow-up (D-F). At both instances, VR pupillometry showed positive RAPD, in the magnitude of 1.0 and 0.8 mm, respectifely; and was also positive. clinically positive. BCVA at presentation was counting fingers at 1,5 m (1.6 logMAR) on right and 1,0 (0 logMAR) on left eye; BCVA on follow-up was 0.04 (1.4logMAR) on tight eye and 1,0 (0 logMAR) on left eye

**Table 2.** Clinical characteristics and results of RAPD testing of patients with optic neuropathy (Group 1).

| ID | Sex | Diagnosis | RAPD assessment with VR pupillometry |  | RAPD assessment with pen light |
| --- | --- | --- | --- | --- | --- |
|  |  |  | RAPA / RAPD [mm] | RAPA / RAPD calculated using only results of the affected eye (RAPAa / RAPDa) [mm] |  |
| 20 | F | ON RE | **1,0\*** | **0,8\*** | **Pos.** |
| 21 | F | ON RE | **1,0\*** | **1,0\*** | **Pos.** |
| 21 | F | *ON R (2 months later)* | **0,8\*** | **0,7\*** | **Pos.** |
| 22 | M | A-AION LE | **0,7\*** | **0,7\*** | **Pos.** |
| 23 | F | ON RE | **0,4\*** | **0,3\*** | **Pos.** |
| 24 | F | A-AION LE | **0,7\*** | **0,8\*** | **Pos.** |
| 25 | M | N-AION RE | **0,5\*** | **0,4\*** | **Pos.** |
| 26 | F | ON LE | 0,1 | 0,1 | Neg. |
| 27 | F | N-AION BE | 0,1 | 0,0 | NRD |
| 28 | F | Optic nerve meningeoma LE | **0,9\*** | **0,9\*** | **Pos.** |
| 29 | M | A-AION LE | **0,5\*** | **0,5\*** | **Pos.** |
| 30 | F | Mycotic optic neuropathy RE | **0,7\*** | **0,5\*** | **Pos.** |
| 31 | F | ON RE | 0,1 | 0,1 | NRD |
| 32 | F | A-AION LE | **1,0\*** | **0,9\*** | **Pos.** |
| 33 | F | Optic nerve meningioma LE | **0,5\*** | **0,5\*** | **Pos.** |
| 34 | F | ON D | **0,7\*** | **0,6\*** | **Pos.** |
| 34 | M | A-AION L | **0,9\*** | **0,8\*** | **Pos.** |
| 35 | F | Status post pituitary tumor surgery | 0,1 | 0,1 | Neg. |
| 36 | F | ON L | **0,6\*** | **0,5\*** | **Pos.** |
| 37 | M | ON D | **0,9\*** | **0,9\*** | **Pos.** |
| 38 | F | ON D | 0,2 | 0,2 | NRD |
| 39 | M | ON D | **1,9\*** | **1,9\*** | **Pos.** |

A-AION = arteritic anterior ischemic optic neuropathy, N-AION = non-arteritic optic neuropathy, ON = optic neuritis, R = right, L = left, \* = positive RAPD on VR pupillometry (difference >0.2 mm), NRD = not reliably determinable.

Figures 6 and 7 show correlations between RAPA and the inter-eye difference in BCVA and MD, respectively. RAPA was statistically significantly associated with the inter-eye difference in visual acuity (Pearson correlation, R = 0.58, p < 0.01) as well as the inter-eye difference in visual-field defect (Pearson correlation, R = 0.68, p < 0.001). All patients with inter-eye MD difference above 5.8 dB had positive RAPD.

**Figure 6.**
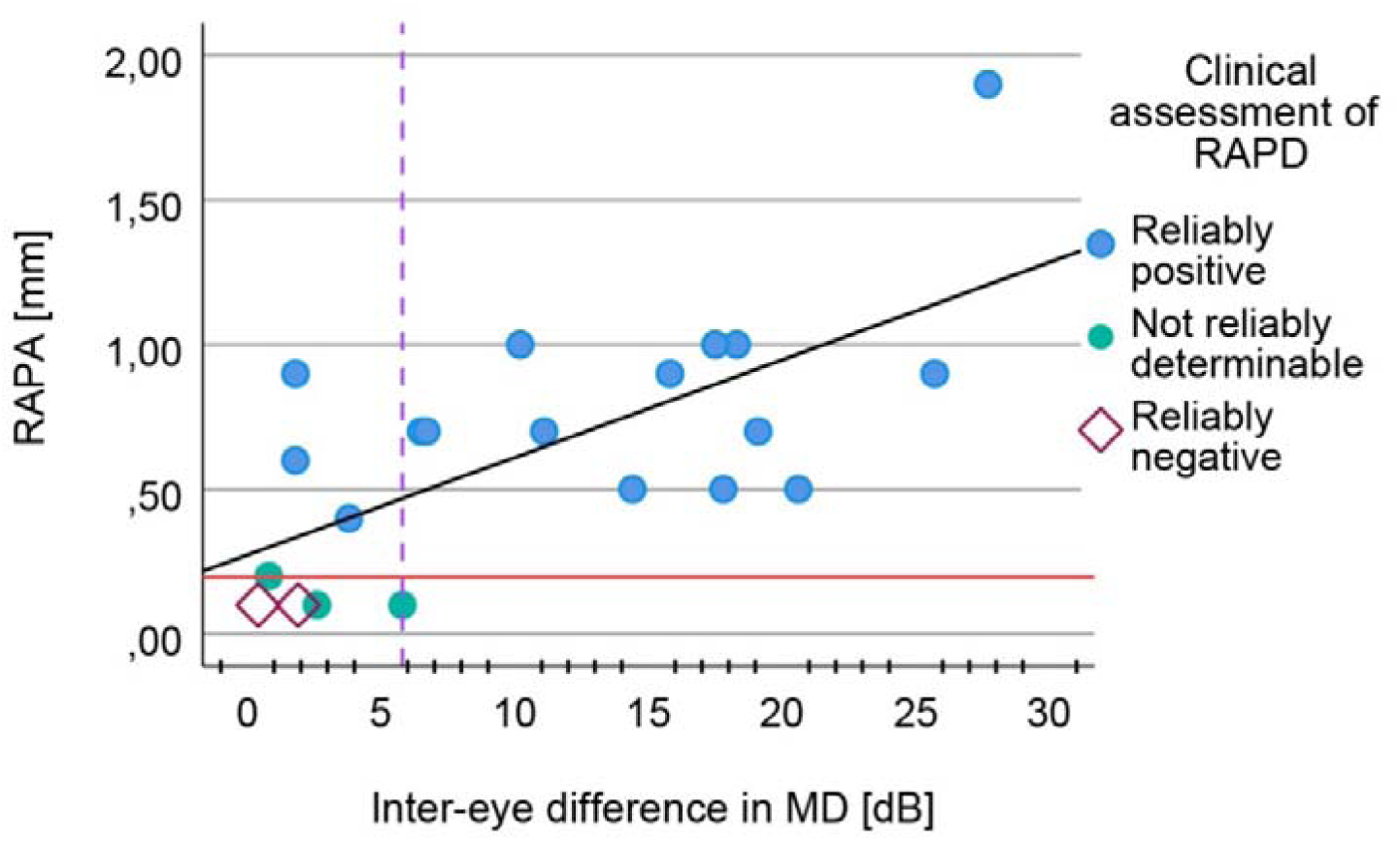
Correlation between inter-eye difference in BCVA acuity and the degree of relative afferent pupillary asymmetry (RAPA). The horizontal line shows the threshold for pathological asymmetry (RAPD), determined as the highest value of controls. Patients are marked with different symbols according to the clinical RAPD measurement (legend on the right). The orange arrow marks a patient with pronounced bilateral visual loss due to A-AION with slightly more preserved function in the left eye (BCVA light perception, 2.8 logMAR on the right and 0.06, 1.2 logMAR on the left). Some patients had the same inter-eye difference and RAPA thus overlapping on the chart.

**Figure 7.**
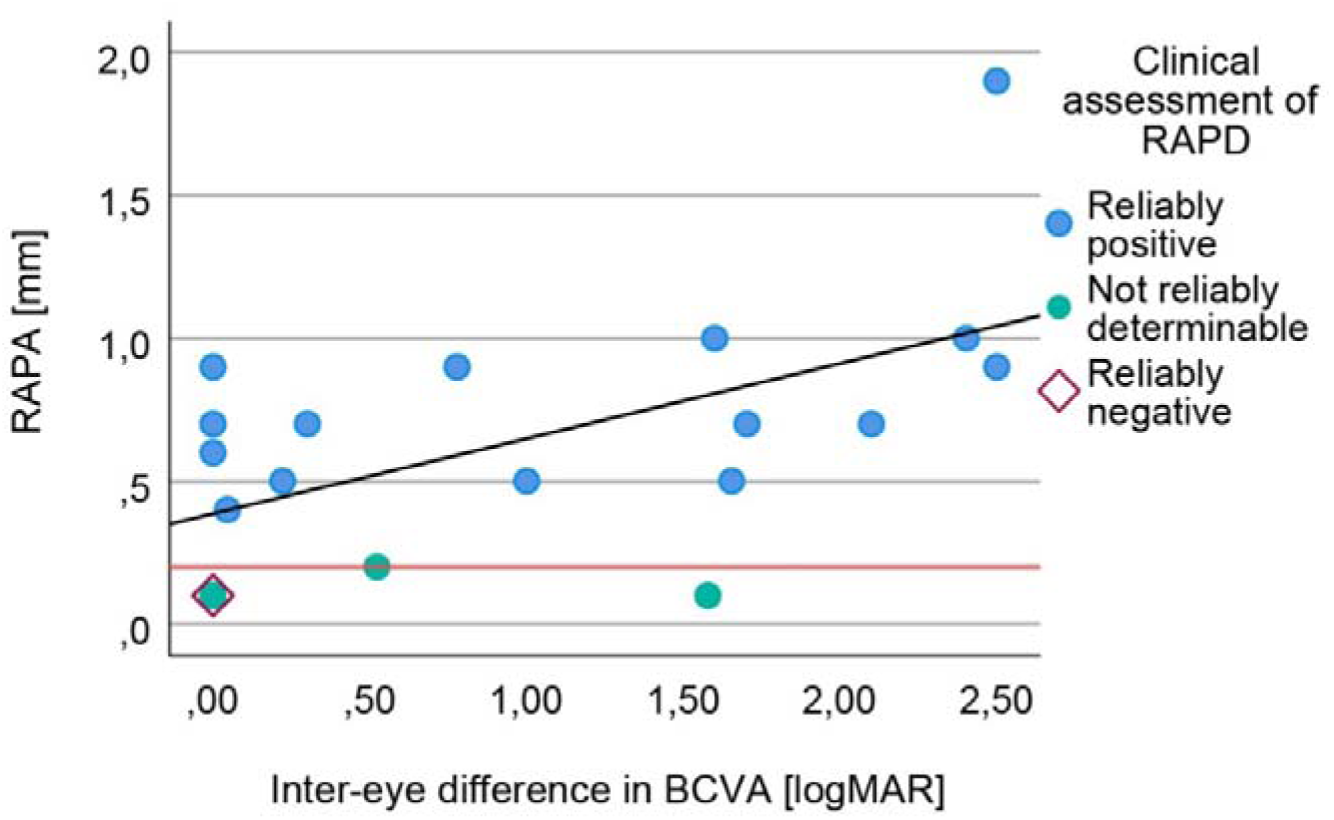
Correlation between inter-eye difference in MD on visual field and the degree of relative afferent pupillary asymmetry (RAPA). The horizontal line shows the threshold for pathological asymmetry (RAPD), determined as the highest value of controls. Patients are marked with different symbols according to the clinical RAPD measurement (legend on the right). Vertical line marks the inter-eye MD difference of 5.8 dB, above which RAPD was positive in all patients. Some patients had the same inter-eye difference and RAPA thus overlapping on the chart.

The results of the evaluation of pupil asymmetry are shown in Table 3. In the control group, VR pupillometry revealed pupillary asymmetry in at least one condition in 84% (16/19) and in both lighting conditions in 74% (14/19). Asymmetry (including anisocoria) was more frequent in darkness. Of the 16 participants in whom asymmetry was detected in at least one lighting condition, all had asymmetry in darkness; 2 only in darkness, and the remaining 14 in both conditions. No participant had asymmetry only in light. Anisocoria (≥0,4 mm) in at least one lighting condition was present in 37% (7/19) controls and in both conditions in 5% (1/19). It was also more frequent in darkness. 7/19 (37%) had anisocoria in dark vs. 1/19 (5%) in light. In all seven, the difference was greater in darkness, by a mean of 0.2 mm (range, 0.1– 0.3 mm).

**Table 3.** Pupil asymmetry evaluation in controls patients with suspected pathological anisocoria.

| ID | Sex | Clinical data | Pupil asymmetry |  |  |  |
| --- | --- | --- | --- | --- | --- | --- |
|  |  |  | VR pupillometry [mm] |  | Measurement on photographs [mm] |  |
|  |  |  | Light | Darkness | Light | Dim light |
| 1 | F | / | 0,2 | 0,3 | - | - |
| 2 | M | / | 0,0 | 0,0 | - | - |
| 3 | F | / | 0,0 | 0,0 | 0,2 | 0,1 |
| 4 | F | / | 0,2 | **0,4** | 0,0 | 0,3 |
| 5 | F | / | 0,0 | 0,0 | 0,2 | - |
| 6 | F | / | 0,2 | **0,4** | - | - |
| 7 | F | / | 0,3 | **0,4** | 0,0 | 0,0 |
| 8 | F | / | 0,0 | 0,2 | 0,2 | 0,0 |
| 9 | M | / | 0,2 | 0,1 | 0,1 | 0,3 |
| 10 | M | / | **0,5** | **0,8** | **0,5** | - |
| 11 | F | / | 0,1 | 0,1 | - | - |
| 12 | F | / | 0,1 | 0,1 | 0,1 | - |
| 13 | F | / | 0,1 | **0,4** | 0,1 | - |
| 14 | F | / | 0,2 | **0,4** | 0,2 | - |
| 15 | F | / | 0,1 | 0,1 | **0,6** | **0,8** |
| 16 | M | / | 0,3 | **0,5** | **0,5** | **0,5** |
| 17 | M | / | 0,0 | 0,3 | - | - |
| 18 | F | / | 0,1 | 0,2 | 0,0 | 0,3 |
| 19 | F | / | 0,1 | 0,3 | 0,3 | 0,1 |
| 40 | M | Larger left pupil, unknown cause | 0,1 | **0,4** | 0,2 | - |
| 41* | F | Congenital right-sided Horner syndrome | **0,4** | **1,1\*** | **0,8** | - |
| 42* | M | Narrower and nonreactive right pupil, unknown cause | **0,4** | **1,3\*** | **0,5** | **0,7** |
| 43 | F | Alternating anisocoria | 0,2 | 0,3 | 0,2 | **0,5** |
| 44 | F | Right pupil transiently narrower, unknown cause | 0,1 | 0,1 | 0,1 | - |
| 45* | M | Left pupil wider and nonreactive, | **1,7\*** | **1,1\*** | **2,2\*** | - |
| 46* | M | Anisocoria, unknown cause | **0,4** | **1,0\*** | - | - |
| 47* | M | Right-sided Horner syndrome after carotid artery dissection | **0,4** | **1,1\*** | **0,6** | **0,8** |

Anisocoria (Asymmetry $\geq 0.4$ mm) is marked in bold, \* = pathological anisocoria ( $>0.8$ mm and/or $>0.3$ mm difference in degree of asymmetry in light and dark), - = measurement not possible.

The largest inter-eye asymmetry of the controls in any light condition was 0.8 mm, and the largest difference in the degree of asymmetry in light and dark was 0.3 mm. Therefore, pathological anisocoria was defined as an inter-eye asymmetry in at least one condition >0.8 mm and/or a presence of difference in the degree of asymmetry between light and darkness >0.3 mm. The upper value (0.8 mm) was consistent with previous observations that physiological anisocoria rarely exceeds 0.8 mm (Lam, Thompson & Corbett 1987).

VR pupillometry confirmed pathological anisocoria in 5/8 patients with suspected pathological anisocoria. Of these, anisocoria could be confirmed by pupil measurement on photographs in only one. In the remaining four, the magnitude of asymmetry on photographs either did not exceed the 0.8 mm threshold (N = 2) or measurement was not feasible (N = 3). Measurement of pupils on photographs was difficult in 22% (6/27) in light and in 60% (16/27) in dim light. The main reasons for difficult measurement were corneal light reflex (19%, 5/27), dark iris color (4%, 1/27), more difficult assessment in dim light (30%, 8/27), and upper-eyelid ptosis (11%, 3/27). Under both lighting conditions, measurement was possible in 41% (11/27) of participants.

### Horner syndrome

There were 2 patients with right-sided Horner syndrome due to a known cause, patient 41 with congenital Horner syndrome and 47 with carotid artery dissection (Table 3). In both, anisocoria was detected by measurements on photographs but did not exceed the pathological threshold. For patient 41, reliable measurement was feasible only in light. In both, VR pupillometry confirmed pathological anisocoria in both, substantially greater in dark (1.1 mm) than in light (0.4 mm) for both (example in Figure 8).

**Figure 8.**
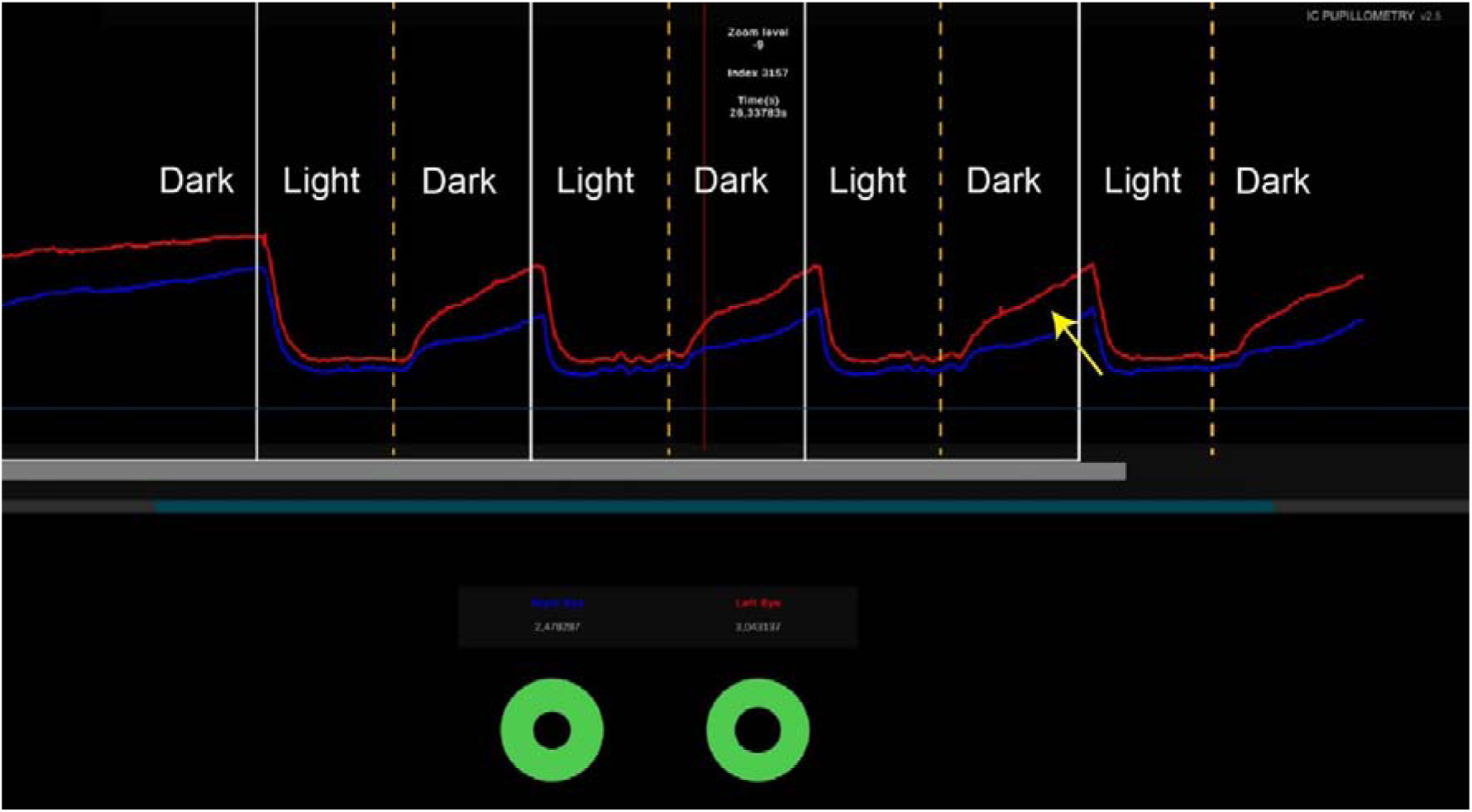
VR pupillometry of patient 47 with right-sided Horner syndrome. A larger difference in pupil size (yellow arrow) is visible in dark when normal pupil dilates. Note qualitative difference in pupil dynamics - delayed dilation of the affected pupil. Vertical white line shows the time points when the light was switched on, and the vertical dashed orange line shows the time points when the light was switched off.

Six participants in Group II had suspected pathological anisocoria without a reliably defined cause. For three of those (40, 43 and 44) VR pupillometry revealed anisocoria within the variability of healthy participants and was concluded to be physiological (example in Fig. 9). For the other three (42, 45 and 46) VR pupillometry revealed pathological anisocoria, in all with the largest asymmetry in darkness (1.0–1.3 mm). None had unilateral ptosis or other pathology indicating Horner syndrome. In two patients, the clinical exam suggested nonreactive pupil on one side, which was also revealed on VR traces (example in Fig. 10).

**Figure 9.**
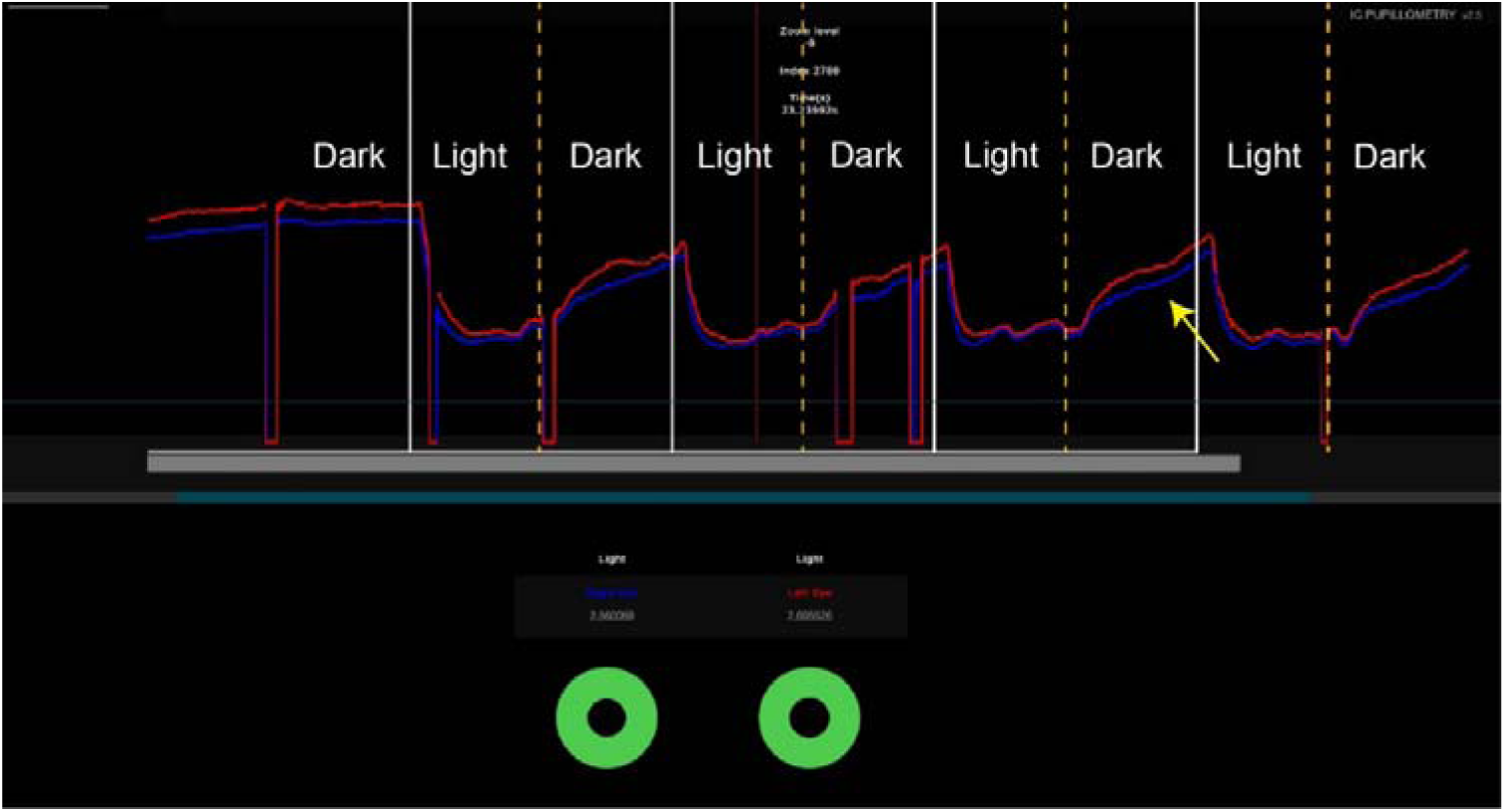
VR pupillometry of participant 43, evaluated for suspected pathological anisocoria. Traces show anisocoria with a wider left pupil (red curve), more pronounced in dark (yellow arrow), but the values did not meet the criterion for pathological anisocoria. Vertical white lines show the time points when the light was switched on, and vertical orange dashed lines show the moments when the light was switched off.

**Figure 10.**
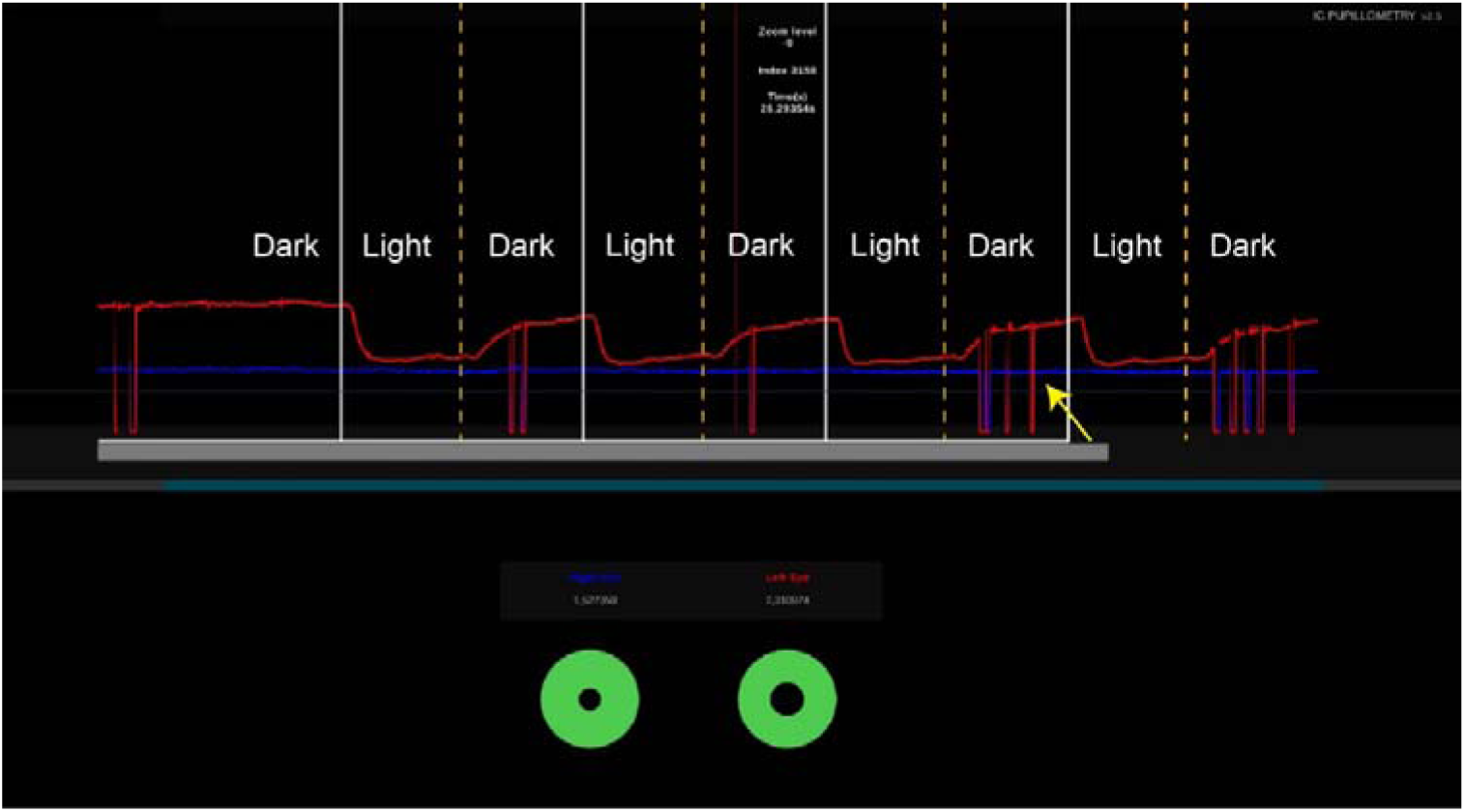
VR pupillometry of participant 42, evaluated for suspected pathological anisocoria. Traces show anisocoria with a wider right (blue curve) and narrower left pupil (red curve) more pronounced in dark (yellow arrow). The longitudinal VR recording shows absent dynamics (nonreactivity) of the right pupil during alternation of light and dark. Vertical white lines show the time points when the light was switched on, and vertical orange dashed lines show the moments when the light was switched off.

## Discussion

We confirmed the utility of HP Reverb G2 Omnicept VR headset and IC pupillometry software (in short, VR pupillometry) for objective pupil measurements. In comparison to conventional testing, VR pupillometry matched detection rate of RAPD and surpassed detection rate of pathological anisocoria. Results support previous studies using (Bruegger et al. 2023, Disse et al. 2024, Negi et al. 2024, Park et al. 2026), and extend the range of devices suitable for pupillary examination.

VR pupillometry identified positive RAPD in all patients with a clinically definite positive RAPD and in none with clinically definite negative or indeterminate RAPD. Previous studies have likewise demonstrated good sensitivity of VR and other automated pupillometry methods for detecting RAPD (Bruegger et al. 2023, Lagrèze & Kardon 1998, Negi et al. 2024, Park et al. 2026). Bruegger et al. detected RAPD with an accuracy of 84% using VR pupillometry, compared with 88% using the flashlight test. In our study RAPD was detected with a similar accuracy of 76% (16/21) in patients with asymmetric optic neuropathy. The limitation preventing VR pupillometry from achieving higher sensitivity and identifying subclinical RAPD does not lie in limitation of the measurement technique, but in the substantial physiological variability of relative afferent pupillary asymmetry (RAPA) (Bruegger et al. 2023, Kawasaki, Moore & Kardon 1996, Negi et al. 2024), resulting in difficulty in defining pathological threshold. In a three-year longitudinal study on healthy participants uising filters of neutral density, 47% showed consistent asymmetry in the same eye (Kawasaki, Moore & Kardon 1996). VR pupillometry revealed even higher percentage of asymmetry, with the majority (84%) of healthy participants exhibiting a small degree of RAPA (0.1 to 0.2 mm). The same range was observed in 5/21 patients with asymmetric optic neuropathy, therefore pupillometry alone could not classify them as having pathological (RAPD). Cohen et al., which compared automated pupillometry with RAPD assessment using neutral-density filters, concluded that automated pupillometry is most useful for identifying and quantifying RAPD in patients with moderate RAPD (Cohen et al. 2015). In our study, a positive RAPD required an interocular difference in MD >5.8 dB. Similarly, a study of RAPD using neutral-density filters identified a threshold of 8.7 dB (Johnson, Hill & Bartholomew 1988). VR Swinging light test offers standardized illumination protocol, simultaneous measurement of the illuminated and non-illuminated pupil, precise quantification of their responses and repeatable testing. This creates opportunities for its use in research setting, for example to monitor changes in patients receiving treatment, where degree of RAPA (even if sub-pathological threshold) could serve as an objective indicator of change in visual function. We demonstrated such monitoring in a patient with optic neuritis (Fig. 5). We found a statistically significant correlation between the degree of RAPA(RAPD) and both visual acuity and visual field, showing that degree of RAPD correlates with conventional parameters of visual perception. Previous studies have reported correlations between RAPD and visual field in patients with optic neuropathy (Bruegger et al. 2023, Johnson, Hill & Bartholomew 1988, Lagrèze & Kardon 1998, Witthayaweerasak, Lertjittham & Aui-aree 2022), although only some with automated infrared pupillometry (Bruegger et al. 2023, Lagrèze & Kardon 1998). An association between visual acuity and RAPD severity measured by infrared pupillometry was found by Siebald et al., Bruegger et al, and Gracitelli et al. (Bruegger et al. 2023, Gracitelli et al. 2016, Siebald et al. 2025). A study of patients with asymmetric glaucomatous damage found a stronger correlation of RAPD with visual-field measures than with parameters of structural ganglion-cell damage (Gracitelli et al. 2016). In addition to identifying RAPD in asymmetric afferent impairment, VR pupillometry can also detect and quantify APD in symmetric impairment. In patients with bilateral impairment and a negative RAPD, the dynamics of the VR pupillometry curve may still reveal severe bilateral APD (Figure 4, bilateral AION). Here too, interpretation is limited by physiological variability and other factors affecting pupillary responses, such as attention and accommodation (Mathôt n.d.). Clinical or research application of APD measurement would require normative data from further studies accounting for all these factors.

VR pupillometry was substantially more sensitive than conventional exam for identifying pupillary asymmetry, detecting pathological anisocoria in 5 vs. 1 of 8 patients, respectively. The increased sensitivity is most likely due to ability to provide precise measurements in darkness, whereas this is impossible with a naked eye and can be falsely negative in dim light, where pupils do not reach maximal dilation. The use of automated pupillometry has been reported relatively frequently to assess RAPD, however only a few studies have used it to measure anisocoria; especially using binocular VR pupillometry. Some studies used the method of recording both pupils in darkness with an infrared camera and measuring pupil size from the recording (Bremner & Smith 2008, Kılınç Hekimsoy et al. 2022, Krzizok, Gräf & Kraus 1995). Some studies reported prevalence of interocular differences of pupil diameters measured using monocular pupillometers in intensive care (Couret et al. 2016). An important limitation of monocular measurement is that it does not provide symmetrical illumination and does not exclude the influence of asymmetric afferent visual impairment (RAPD). Reliable assessment of anisocoria requires symmetrical illumination and simultaneous measurement of both pupils.

Longitudinal VR recordings revealed qualitative abnormalities in pupillary responses. These may be diagnostically relevant for determining the cause of anisocoria and identifying pathological responses even when interocular asymmetry is small, which could be investigated in further studies on larger samples with diverse, established causes of anisocoria. The utility of qualitative pupillary-response analysis has been examined in a small study of patients with Horner syndrome (Disse et al. 2024). In our study, VR pupillometry also identified pathological anisocoria in both patients with known Horner syndrome. Qualitative analysis showed dilation lag of the affected pupil but otherwise relatively preserved light-response dynamics (Figure 8). Horner syndrome poses a particular diagnostic challenge in clinics because it is most pronounced in darkness, when reliable measurement of inter-eye pupillary asymmetry is difficult. It is conventionally confirmed with cocaine or apraclonidine testing, but these tests are time-consuming and may be unavailable because of supply or other constraints. Larger studies could establish whether VR pupillometry could reliably confirm Horner syndrome without the need of pharmacological testing. In two other patients in whom asymmetry was also greater in darkness, qualitative assessment showed absent pupillary reactivity on one side, i.e. tonic pupil (Fig. 10). This could help in identifying the cause of anisocoria, such as Adie’s pupil (Bremner & Smith 2007, Kelbsch et al. 2019, Thompson 1977, Thompson 1984).

VR pupillometry of healthy participants provides important insights into the nature of physiological variability of pupillary behavior. Classical definition of physiological anisocoria is a small inter-eye difference (usually approximately 1 mm), equal in light and dim conditions (‘Anisocoria - EyeWiki’ n.d.). Only a few recent studies have reported that it is more frequent and/or more pronounced in lower illumination. Using infrared video pupillometry, Hekimsoy et al. found anisocoria in 17% participants under low-mesopic conditions (0.1 cd/m²) vs. 3% under high-photopic conditions (100 cd/m²) (Kılınç Hekimsoy et al. 2022). Our VR pupillometry findings support these observations, detecting anisocoria in 37% in darkness vs. 5% under light conditions (150 cd/m²). All participants with anisocoria had, on average, a 0.2-mm greater difference in darkness. The finding that physiological anisocoria is greater in darkness is clinically important, because awareness of this phenomenon may prevent unnecessary referrals and diagnostic tests. Hekimsoy et al. noted that defining anisocoria as an absolute interocular difference in pupil diameter of at least 0.4 mm may bias the analysis. When they applied a criterion of a 6% difference in diameter or 15% in pupil area, the difference in anisocoria prevalence between lighting conditions diminished (20% under mesopic vs. 16% under photopic illumination) (Kılınç Hekimsoy et al. 2022). The absolute-difference criterion is nevertheless more clinically relevant because it more closely reflects unaided visual assessment.

### Strengths and limitations of the study and proposals for improving VR pupillometry

A strength of our study is the use of binocular pupillometry, which, unlike monocular pupillometry, permits bilateral darkening, symmetrical illumination, and simultaneous measurement of both pupils’ responses to direct and consensual stimulation. Objectivity is the principal advantage of VR pupillometry over conventional flashlight tests, which depend heavily on examiner experience and ambient lighting. VR pupillometry may provide more consistent, unbiased results regardless of examiner experience, thereby improving diagnostic reliability (Park et al. 2026). Ours is the first study to use the same VR-headset model to investigate both anisocoria and RAPD. We obtained longitudinal data on pupillary dynamics during bilateral and alternating illumination, enabling more complex retrospective analyses, including assessment of the effect of anisocoria on RAPD. The study is limited by incomplete age matching between controls and patients and by the relatively small patient samples. Previous studies have shown that age affects pupil diameter, with smaller pupils in older individuals, which may influence anisocoria detection (Lam, Thompson & Corbett 1987). Because controls were relatively younger, they would be expected to have larger pupils and more anisocoria. We established normative thresholds from their highest values, thereby reducing the likelihood of false-positive results. To date, no significant effect of age on pupillary responses used to determine RAPD has been reported (Gracitelli et al. 2016).

For further research, more precise results, and comparison of VR pupillometry using the HP Omnicept and other headsets, measurement units should be standardized or converted to those used in previous studies, particularly for RAPD quantification. We decided to use mm to most closely resemble clinical testing, whereas other studies report RAPD in % (Meethal et al. 2021) or Log values (Pillai et al. 2019, Wilhelm et al. 2001). Some studies also used a different alternating-illumination paradigm, for example with intervening dark intervals, which our protocol did not include. Initial conditions could be standardized further by several minutes of darkness; Wang et al. recommended as much as 20 minutes of dark adaptation (Wang et al. 2015). VR pupillometry offers more accurate assessment of RAPD in patients with anisocoria however the testing protocol has not yet been standardized. RAPD is traditionally determined from the pupillary response of the affected eye. In cases of iris pathology or iatrogenic mydriasis, it can also be assessed indirectly by observing the unaffected eye, the so-called reverse RAPD (Nakamura et al. 2023, Satou et al. 2016). Because VR pupillometry permits simultaneous measurement of both pupils, we attempted to reduce the effect of low-grade anisocoria by averaging the responses of both pupils. This approach has also been used in some studies of bilateral pupillometry (Gracitelli et al. 2016, Wilhelm et al. 2001). In a large degree of anisocoria and/or unilaterally tonic pupil however, only the normally constricting pupil should be used for most reliable RAPD evaluation. VR pupillometry creates new opportunities to investigate pupillary responses in healthy individuals and patients (Omary et al. 2019). Quantifying pupillary responses with VR pupillometry could provide insight into which regions of the retina and optic nerve contribute most strongly to APD. In a patient with optic neuritis, VR pupillometry continued to show a substantial RAPD despite a marked reduction in visual-field loss. This suggests that papillomacular fibers and/or the number of affected fibers are important determinants of RAPD severity. Similar findings have been reported by Gracitelli et al. (Gracitelli et al. 2016). VR headsets can display different fixation targets that may elicit accommodative responses and complex scenes that may stimulate higher brain centers, enabling measurement of their influence on sympathetic tone (McDougal & Gamlin 2015, Xiong et al. 2021). Future VR headsets may incorporate additional protocols for evaluating pupillary health, including pupil shape, dynamics (latency, reaction time, maximum velocity, etc.), post-illumination response characteristics, and chromatic pupillometry.

## Conclusions

We confirmed the usefulness of a new method of VR pupillometry using the HP Reverb G2 Omnicept Edition VR headset and IC Pupillometry software. VR pupillometry achieved the same RAPD detection rate and exceeded the detection rate of pathological anisocoria compared with conventional pupil testing. VR pupillometry may make an important contribution to diagnosis in clinical practice, for example in the neuro-ophthalmology clinic when assessing RAPD and evaluating anisocoria, particularly in suspected Horner syndrome. In addition to its clinical utility, the method also has research potential. Quantitative and qualitative data on the dynamics of pupillary responses may substantially contribute to the objective monitoring of patients in clinical studies and to a deeper understanding of the physiological variability of pupillary responses.

## Data Availability

All data produced in the present study are available upon reasonable request to the authors

